# Clinical characteristics of children with COVID-19 admitted in a tertiary referral center in Perú

**DOI:** 10.1101/2020.09.18.20186866

**Authors:** Christian Chiara Chilet, Medalit Luna Vilchez, Julio Maquera Afaray, Blanca Salazar Mesones, Diana Portillo Alvarez, Ramiro Priale Miranda, Franklin Mendoza, Aldo Munayco, Jenny Baca, Mitsi Santiago, Jose W. López

## Abstract

**Introduction:** COVID-19 pandemic represents a big impact on children’s health, this study describes the behavior of the disease in hospitalized pediatric patients in the Instituto Nacional de Salud del Niño San Borja (INSN-SB).

**Methods:** Retrospective study of patients with confirmed COVID-19 diagnostic between March and July 2020. Demographic, clinic, laboratory, radiology and treatment data were collected and for the analysis descriptive statistics were included.

**Results:** From a total of 91 patients. 36.3% (33) were female. The age group who was affected the most were school children with a median age of 4 years old (IQR 1-8). Patients who came from Lima represented 61.5%. Previous contact was determined in 30.8% of the cases. PCR results for SARS CoV-2 were positive in 50.6% of the cases and 49.4% in the quick tests. Comorbidity was present in 53.8% of the cases. Most frequent symptoms were fever (39.6%), general discomfort (23.1%), cough (19.8%) and shortness of breath (14.3%). Presence of MIS-C was confirmed in 6 patients. Use of antibiotics represented 76.9% of the cases. The most frequent radiology pattern was bilateral interstitial (57.7%). Comorbidities were present in 68.2% (15/22) of patients in PICU. From a total of 9 deceased patients, 6 were admitted in PICU and 8 presented associated comorbidities.

**Conclusions:** COVID-19 in children displays mild and moderate clinical manifestations. A great proportion of patients exhibited comorbidities, especially PICU patients and the ones that died.

**What is known about the subject:** In pediatric patients, the prevalence and severity of COVID-19 are usually low, however, in the presence of MIS-C, greater severity and probability of admission to the PICU is observed.

**What this study adds:**

- This study describes the results of complex pediatric patients and the associated comorbidity in LMIC setting that showed greater severity and admission to the ICU.
- Microbiological isolates in cultures were low, therefore the initiation of empirical antibiotic therapy is not justified in most cases.

## Introduction

The new Coronavirus pandemic is the biggest public health crisis which the world still faces. It has been estimated that 90% of the cases correspond to the adult population and only between the 1% and 5% to the pediatric population (1), from which more than 90% are asymptomatic forms of presentation, mild or moderate and only 5.9% are severe (2), a smaller percentage compared with the severity reported in adults which is from 12.6% to 23.5% (3). It has been reported that from 48 pediatric patients admitted to PICU (pediatric intensive care unit), 40 (83%) had some comorbidity (4). Another study reported that from 651 children, 276 (46%) had at least one comorbidity (6).

Furthermore, the presence of pediatric multisystem inflammatory syndrome (MIS-C) has been documented which can be severe and present itself with significant cardiovascular compromise (5). It has been reported that patients with MIS-C presented 5 times higher probability of being admitted to an ICU (6). In Peru it has been determined that the incident rate is 10 times higher in adults than in children (7) and COVID-19 behavior is unknown in pediatric patients with and without comorbidities.

Peruvian health systems consist of the public and private system, the INSN-SB is a public center that takes care of pediatric patients who have been referred from another national hospital in their majority. Most of our patients are registered in the “Sistema Integral de Salud” (SIS) which is the public insurance created firstly as a partial financing program and to date it has universal coverage for any person who resides in Peru (8).

The aim of the study is to describe the clinic-epidemiological and treatment characteristics of COVID-19 diagnosed children hospitalized in the INSN-SB which characterizes for being a specialized surgery center of high complexity.

## Methods

### Site and study design

The INSN-SB is a surgical and specialized pediatric hospital of high complexity of national reference that attends mainly patients with cardiovascular surgical pathology, neurosurgery as well as patients with onco-hematologic disease and organ transplant. The INSN-SB has 306 beds from which 24 beds are patients with confirmed COVID-19 diagnosis, 25 beds for suspicious cases and 12 beds for PICU.

A retrospective study reviewing clinical histories of patients with confirmed COVID-19 diagnostic, during March to July 2020 was performed.

### Participants

Hospitalized patients who are younger than 18 years old with confirmed COVID-19 diagnosis were included through serological and molecular tests. Additionally, patients with incomplete data from the clinical history were not included.

For laboratory diagnosis of SARS-CoV-2 infection, tests based on lateral flow immunochromatography and/or molecular tests using nasopharyngeal swabs for polymerase chain reactions (PCR) were performed. Immunochromatography tests were performed and processed in our institution, on the other hand molecular tests samples were taken in our institute but processed in the National health Institute (INS). Moreover, laboratory confirmatory tests were performed in asymptomatic patients previous to chemotherapy or in the surgery perioperative (emergency or elective). Image tests were prescribed by a pediatric doctor and were performed following the guidelines for attendance of COVID-19 patients of the INSN-SB (Figure 1).

**Figure 1.**
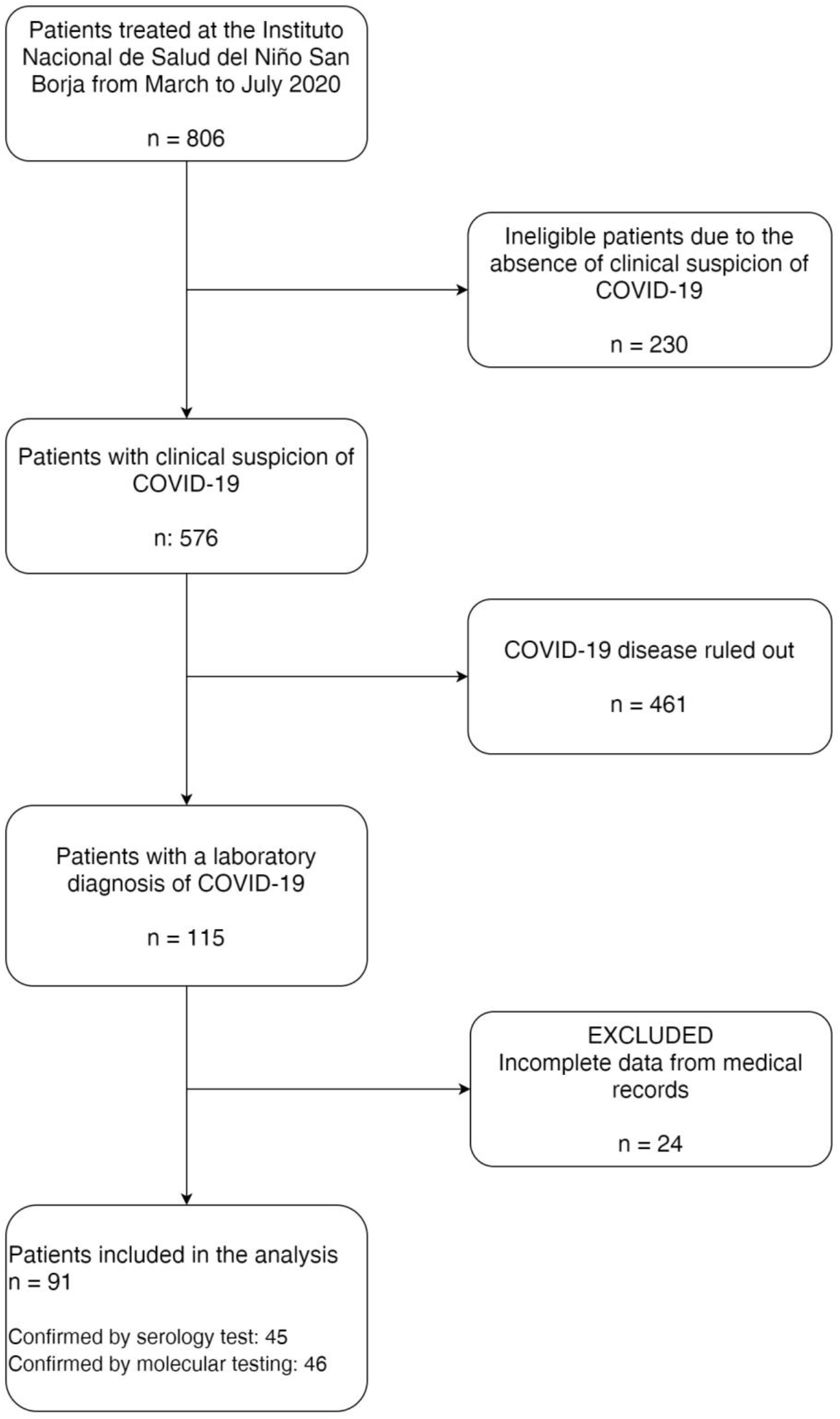
**Flow diagram of the selection of patients with COVID-19 in the Instituto Nacional de Salud del Niño San Borja during the period from March 1st to July 31st, 2020**.

### Variables of interest

COVID-19 cases definition was considered according to the institutional norm of the Peruvian Ministry of health (MINSA) (9), as well as the WHO (World Health Organization) definition of MIS-C which includes children and adolescents between 0 and 19 years old (10).

Evaluated epidemiological variables were sex, age, city of origin, weight, height, comorbidity and direct contact with SARS CoV-2 infected patients. The clinical variables included were: date of symptoms onset and the presence or absence of symptoms like general discomfort, cough, shortness of breath, diarrhea, abdominal pain, nausea, vomit, headache, rhinorrhea and pharyngeal pain. Clinical signs considered were: fever, conjunctival injection, rash, dyspnea, rhonchus, wheezing, crepitant, sub crepitant, convulsions, pharyngeal exudate, pretibial edema or limb peeling and strawberry tongue.

Laboratory tests for SARS CoV-2 infection were both rapid and molecular tests. Furthermore, laboratory test like: hemoglobin, leukocytes, white blood cell differential, platelets, albumin, creatinine, ferritin, C reactive protein (CRP), procalcitonin, erythrocyte sedimentation rate (ESR), lactic dehydrogenase (LDH), hepatic profile, prothrombin time (PT), D-dimer, creatine phosphokinase (CPK), creatine phosphokinase-MB (CPK-MB) and lactic acid were included.

As microbiological variables, IgM antibodies for *Mycoplasma pneumoniae*, Cytomegalovirus, Epstein-Barr virus, influenza A and B, herpes 1 and parvovirus B19 were requested. Additionally, blood cultures were performed. Furthermore, radiology patterns in thorax radiology were described.

Treatment received during COVID-19 infection was described based on groups of drugs. (antibiotics, antifungal, antiviral, antiparasitic, anticoagulant, antipyretic, human immunoglobulin, corticosteroids and vasoactive substances).

Finally, clinical picture severity, PICU requirement and date of admission were evaluated. In addition, MIS-C presence or absence, as well as hospital stay considering final patient condition and the cut-off point as the last day of July 2020.

### Data analysis

The study data were collected and administered using the Integrated Hospital Management System (SIS Galen Plus) (11) and the electronic data capture tools REDCap (12,13) housed in the INSN-SB.

The statistical analysis used the STATA version 16 program (StataCorp. 2019. Stata Statistical Software: Release 16. College Station, TX: StataCorp LLC). In the first place, descriptive statistics of the variables of interest were performed, where the quantitative variables were represented by the median and the interquartile range, due to the non-normal distribution of the values, and the qualitative variables were analyzed using frequency tables.

### Ethical aspects

The present study was approved by the Institutional ethics committee in research of the INSN-SB with the research code PI-429.

### Patient and Public Involvement

The patients were not involved in the development of the research question and outcome measures.

## Results

Between March and July 2020, 576 of patients were admitted to the INSN-SB with COVID-19 suspicion, from which the 19.9% (n=115) had confirmed diagnosis by molecular or quick tests. A total of 91 patients were analyzed. Most patients were male, the prevalent age group was schoolchildren with a median of 4 years old (1-8). Only 2 neonates patients were reported, 61.5% (n=56) of patients originated from Lima and most of them 89.1% (n=81) had incomplete vaccines. From patients that needed a surgical procedure, 53.8% were an emergency.

Comorbidities were present in 53.8% (n=49) of patients, being cardiovascular and neurological the most frequent.

Admitted to PICU were 22 patients, mostly male (54.5%) with an age median of 2.5 years old (1-9). No patient presented complete vaccines and the majority (66.7%) were subject to elective surgery. It was not possible to determine if patients had a known contact with COVID-19 carriers in 81.8% of the cases. Underlying comorbidities were found in 68.2% of patients (Table 6). Regarding imaging studies, 61.9% (n=13) of patients presented abnormal thorax radiography. Deceased patients represented 27.3% (n=6/22).

**Table 1.** Demographic characteristics of COVID-19 patients treated in the INSN-SB (n=91).

|  |  |  |  |
| --- | --- | --- | --- |
| Characteristics |  | Total |  |
|  |  | n | % |
| Sex |  |  |  |
|  | Feminine | 33 | 36,3 |
|  | Masculine | 58 | 63,7 |
| Age (years) |  | 4 (1-8) |  |
| Origin |  |  |  |
|  | Lima | 56 | 61,5 |
|  | Province | 35 | 38,5 |
| Incomplete vaccination |  | 81 | 89,1 |
| Admitted due to surgery |  |  |  |
|  | Emergency | 21 | 53,8 |
|  | Elective | 18 | 46,2 |
| Contact |  |  |  |
|  | Family | 24 | 26,4 |
|  | Patient | 4 | 4.4 |
|  | Unknown | 63 | 69.2 |
| Diagnostic test |  |  |  |
|  | Quick test | 45 | 49,4 |
|  | Molecular test | 46 | 50,6 |
| Radiography |  |  |  |
|  | Normal | 45 | 63,4 |
|  | Abnormal | 26 | 36,6 |
| Comorbidity presence |  |  |  |
|  | Yes | 49 | 53,8 |
|  | No | 42 | 46,2 |
| MIS-C presence |  |  |  |
|  | Yes | 6 | 6.6 |
|  | No | 85 | 93.4 |
| Final patient's state |  |  |  |
|  | Survivor | 82 | 90.1 |
|  | Non-survivor | 9 | 9.9 |
| *Median IQR |  |  |  |

**Table 2.** Clinical and radiological characteristics of COVID-19 patients treated in the INSN-SB (n=91).

| Characteristics |  | Total |  |
| --- | --- | --- | --- |
|  |  | n | % |
| Symptoms |  |  |  |
|  | General discomfort | 21 | 23,1 |
|  | Cough | 18 | 19,8 |
|  | Shortness of breath | 13 | 14,3 |
|  | Vomit | 11 | 11,2 |
|  | Abdominal pain | 10 | 11,0 |
|  | Rhinorrhea | 8 | 8,8 |
|  | Nausea | 6 | 6,6 |
|  | Diarrhea | 3 | 3,3 |
|  | Pharyngeal pain | 3 | 3,3 |
|  | Convulsions | 2 | 2,2 |
|  | Headaches | 2 | 2,2 |
| Signs |  |  |  |
|  | Fever (Temp in °C) | 36 | 39,6 |
|  | Dyspnea | 12 | 13,2 |
|  | Rhonchus | 9 | 9,9 |
|  | Wheezing | 5 | 5,5 |
|  | Convulsions | 5 | 5,5 |
|  | Crackles | 4 | 4,4 |
|  | Lymphadenopathy | 4 | 4,4 |
|  | Rash | 3 | 3,3 |
|  | Pretibial edema | 2 | 2,2 |
|  | Strawberry tongue | 1 | 1,1 |
|  | Conjunctival injection | 1 | 1,1 |
| Radiological thorax pattern |  |  |  |
|  | Bilateral interstitial | 15 | 57,7 |
|  | Unilateral alveolar | 8 | 30,7 |
|  | Unilateral interstitial | 3 | 11,5 |
|  | Bilateral alveolar | 2 | 7,6 |

**Table 3.** Laboratorial and microbiological characteristic of COVID-19 patients treated in the INSN-SB.

| Characteristics |  | Median | Interquartile range |
| --- | --- | --- | --- |
| Hematology |  |  |  |
|  | Hemoglobin (g/dL) | 11,8 | 9,8-12,8 |
| | Leukocytes ($10^3$/uL) | 10,8 | 7,43-14,57 |
| | Lymphocytes ($10^3$/uL) | 3,04 | 1,82 – 4,91 |
| | Monocytes ($10^3$/uL) | 0,63 | 0,45 – 0,96 |
| | Neutrophils ($10^3$/uL) | 5,05 | 3, 10-9,24 |
| | Band neutrophils ($10^3$/uL) | 0 | 0,0 |
| | Platelets ($10^3$/uL) | 314 | 233,0-399,5 |
| Biochemistry |  |  |  |
|  | C-Reactive protein (mg/L) | 9,4 | 2,1-96,7 |
|  | Erythrocyte sedimentation rate (mm/h) | 28 | 15-50 |
|  | Creatinine (mg/dL) | 0,33 | 0,22-0,45 |
|  | Albumin (g/dL) | 4,01 | 3,49-4,5 |
|  | Aspartate transaminase (AST) (U/L) | 31 | 22-43 |
|  | Alanine transaminase (ALT) (U/L) | 17 | 13-32 |
|  | Ferritin (ng/dL) | 276,10 | 93,8-468,1 |
|  | Total bilirubin (mg/dL) | 0,62 | 0,34-1,22 |
|  | Prothrombin time (sec) | 14,8 | 13,7-16,2 |
|  | Fibrinogen (mg/dL) | 344,5 | 266-502 |
|  | D-dimer (ug/mL) | 0,69 | 0,27-1,65 |
|  | Procalcitonin (ng/mL) | 0,14 | 0,05-0,65 |
|  | Creatine phosphokinase (CPK) (U/L) | 110 | 40-254 |
|  | CPK-MB (U/L) | 29.9 | 24,5-38 |
|  | Lactic acid (mmol/L) | 2 | 1,4-3,4 |
|  | Lactic dehydrogenase (LDH) (U/L) | 650 | 442-848 |

| Serology | n | % |
| --- | --- | --- |
| *Mycoplasma* | 9 | 9,9 |
| Cytomegalovirus | 3 | 3,3 |
| Epstein-Barr virus | 2 | 2,2 |
| Influenza B | 1 | 1,1 |
| Herpes 1 | 1 | 1,1 |
| Parvovirus B19 | 1 | 1,1 |
| Gram staining | n=21 | % |
| Gram positives | 9 | 42,9 |
| Gram negatives | 8 | 38,1 |
| Fungus | 4 | 19,0 |

**Table 04:**
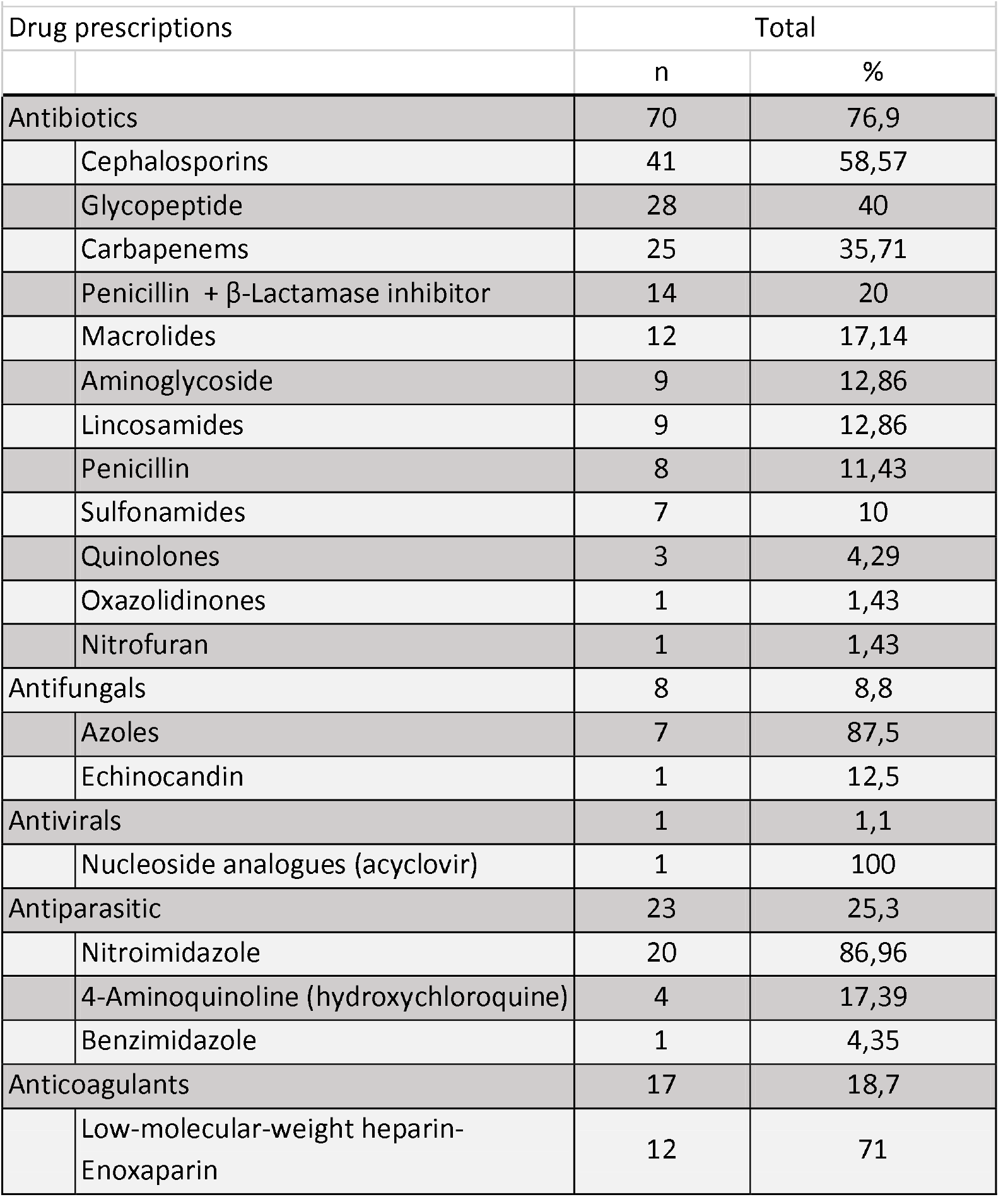
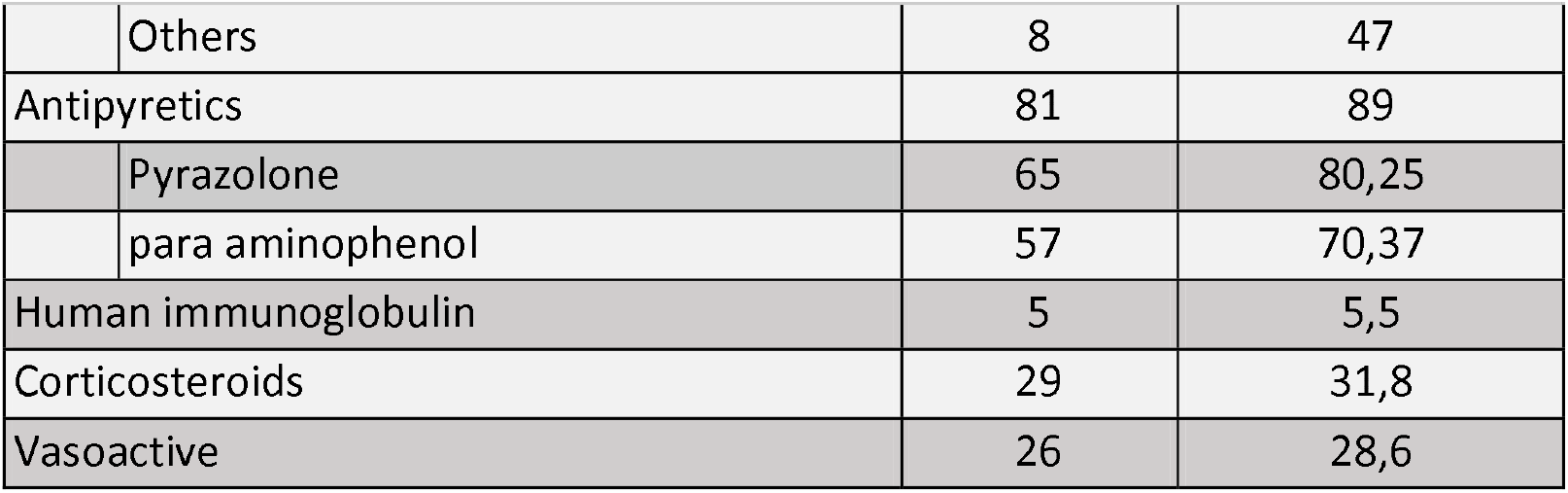
Frequency of prescription in patients with COVID-19 treated in the INSN SB (n=91)

**Table N°5:**
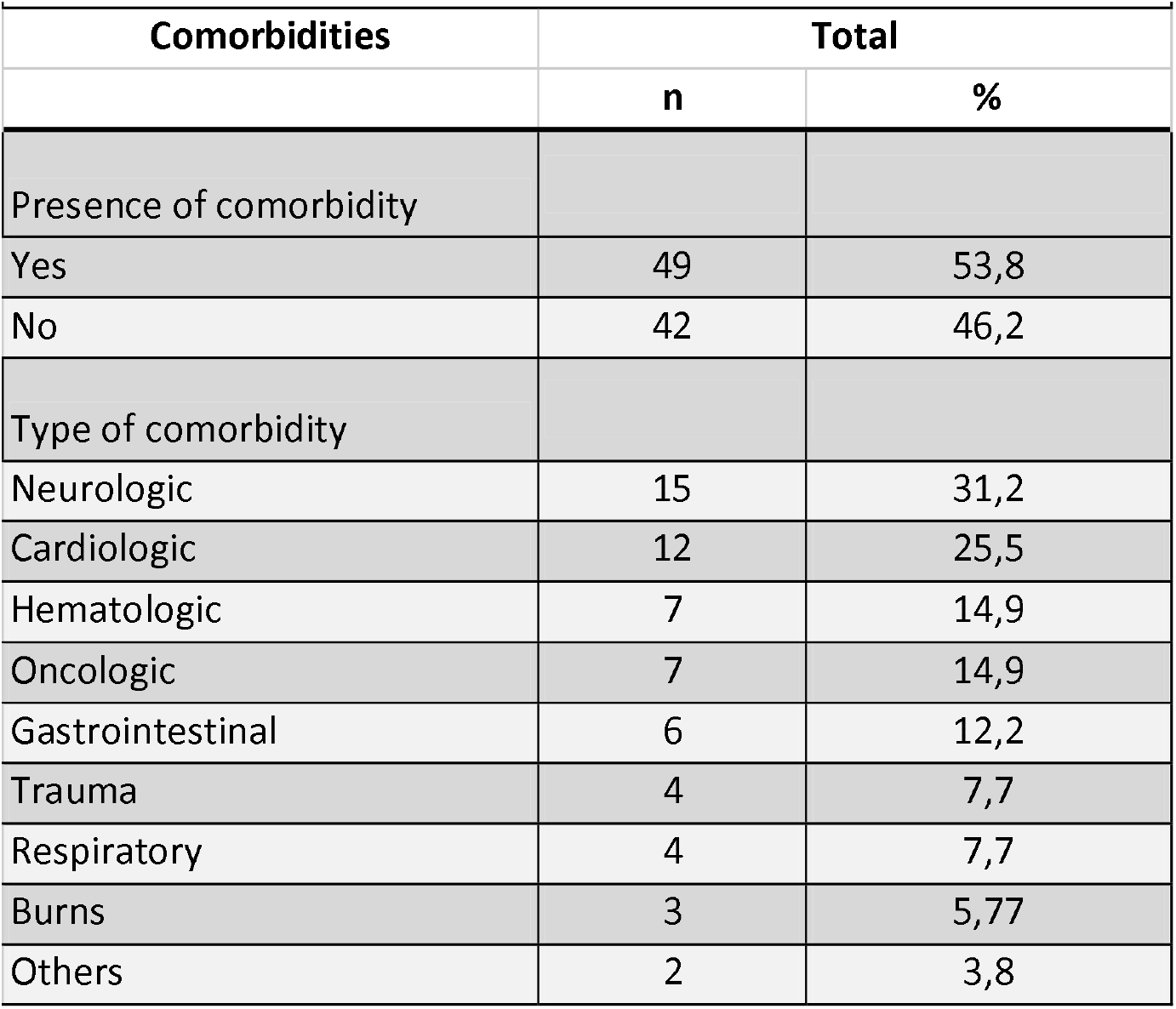
Comorbidities of patients with COVID-19 treated in the INSN-SB(n=91)

**Table 06.**
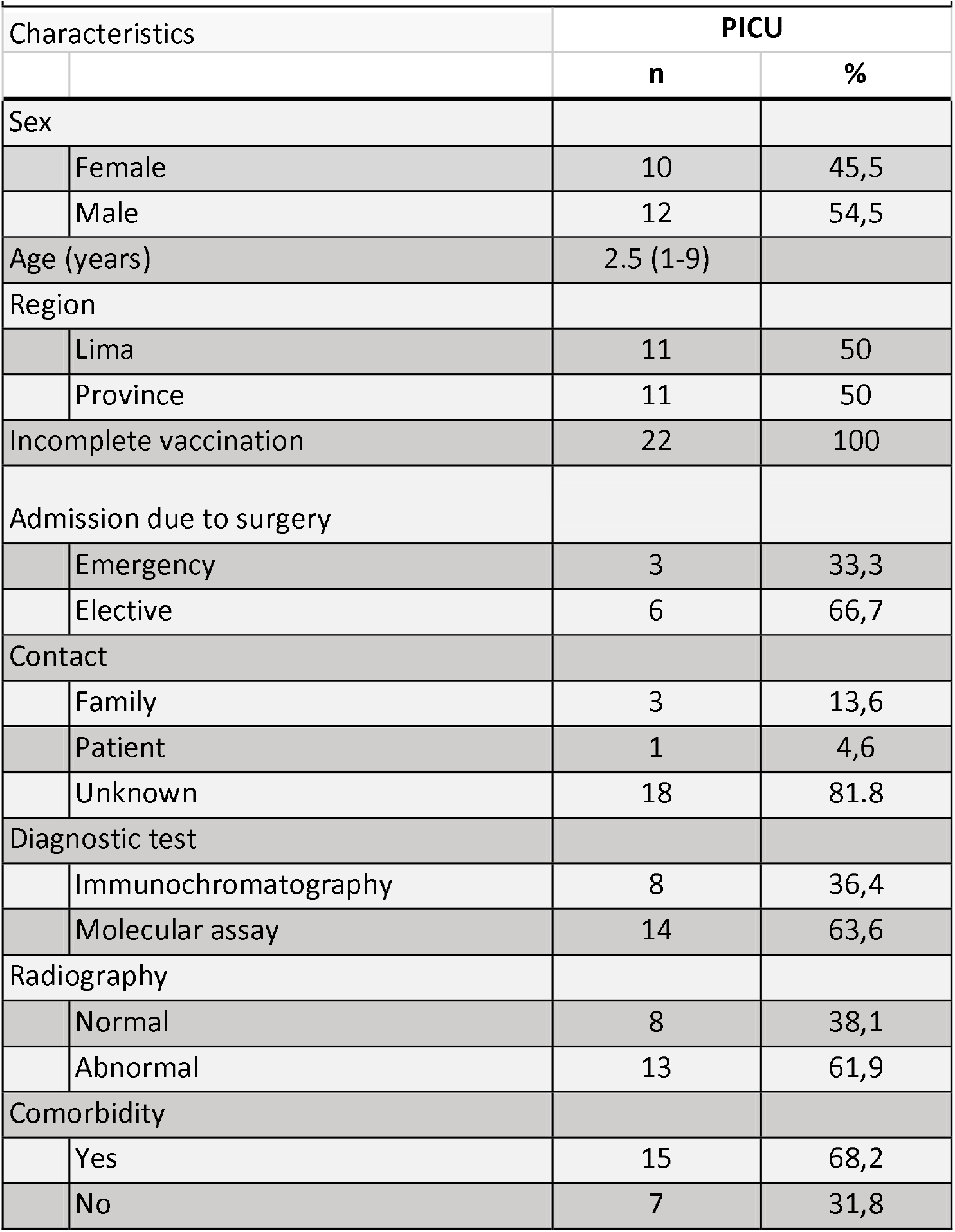
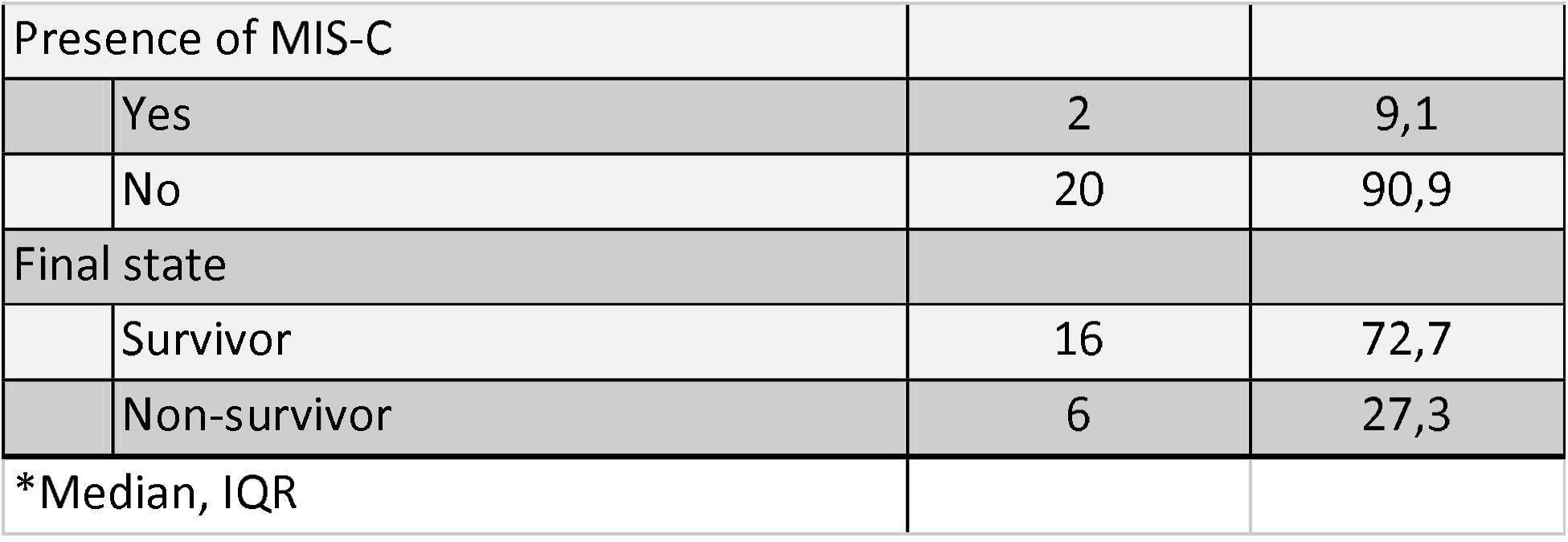
Demographic characteristics of the PICU patients with COVID-19 treated in the INSN-SB(n=22)

The reported symptomatology was: general discomfort 23.1% (n=21), cough 19.8% (n=18), shortness of breath 14.3% (n=13), vomiting 12.1% (n=11). Moreover, as evident signs patients reported: fever 39.6% (n=36), dyspnea 13.2% (n=12), rhonchus 9.9% (n=9) and convulsions 5.5% (n=5).

Overall, most patients had a normal thorax radiography 63.6% (n=14). Additionally, the most frequent thorax radiology pattern was the bilateral interstitial 5737% (n=15).

With regards to the 9 deceased patients, 4 had a positive molecular test and 5 had a positive quick test, age range median was 5 years old (1-12), 88.9% (8/9) of patients presented comorbidities, 1 patient presented both cardiovascular and neurological comorbidities, 2 cardiological, 2 oncological, 1 hematological, 1 gastrointestinal and 1 with a skin burn comorbidity.

In relation to MIS-C, only 6.6% (n=6) of patients presented the criteria exposed by the WHO, of which the majority did not require PICU admission (n=4).

Results from patients who had laboratory tests on admission, showed that the main alterations were the C-reactive protein (CRP) levels with 9.4 mg/L (1.1-96.7) and D-dimer with 0.68 ug/ml (0.27-1.65). There was no sign of greater alteration of procalcitonin with 0.14 (0.005-0.365) nor in the aspartate transaminase (AST), alanine transaminase (ALT) and creatinine levels. Concerning the hemogram, most of our patients were not anemic, hemoglobin 11.8 g/dl (9.8-12.8). No leukocytosis, neutrophilia, left deviation or pronounced lymphocytopenia (leukocytes in 10800 × 103/uL (7430-1450), lymphocytes 3040 × 103/uL (1820 −4910), neutrophils 5045 × 103/uL (3095 – 9240)) (Table 3) were found.

Mycoplasma pneumoniae was the most isolated microorganism with 9.9% (n=9) by IgM serology, followed by Cytomegalovirus with 3.3% (n=3), Epstein-Barr virus with 2.2% (n=2) and Influenza with 1.1% (n=1). By means of blood culture, the most abundant microorganism isolated were Gram positives with 42.9% (n=9) and in less proportion Gram negatives with 38.1% (n=8) and fungus with 19% (n=4).

In regard to the drugs administered for other pathologies, 76.9% (n=70) of patients received antibiotics from which the most prescribed were cephalosporins with 58.6% (n=41) and glycopeptides with 40% (n=28). Moreover, it is important to mention that 40% (n=25) of patients received carbapenem. Macrolides like azithromycin was used in 17.14% (n=12) of the cases. Concerning antifungals, most patients received therapy with azoles 87.50% (n=7). Antiparasitic were prescribed for 25.3% (n=23) of patients and hydroxychloroquine represented the 17.4% (n=4) of these prescriptions. Additionally, antipyretics were used in 89% (n=81) of patients from which the majority received paracetamol, an 18.7% (n=17) of patients received anticoagulants, 31.9% (n=29) received corticosteroid and the 28.6% (n=26) vasoactive. With reference to MIS-C, the 5.5% (n=5) of patients received human immunoglobulin (Table 4).

## Discussion

In this study, it was found that the majority of patients presented a mild clinic picture and almost a third of patients were admitted to PICU in an associated comorbidity context. In addition, a group of patients were described with diagnostic criteria of MIS-C from which all of them evolved in a favorable manner.

Similar to the findings reported in our study, the Center for Disease Control and Prevention found that from 576 hospitalized children the median age was 8 years old (14) and in a systematic review the age mean was 8.9 years old (15) and males were predominant (14,15). Furthermore, only a third of patients presented family contact (26.4%), in contrast with other studies where they showed family contact as a background in more than 40% of cases (15,16), this finding can be explained due to the lack of information about diagnosis in the family surroundings.

Even though in this study respiratory symptoms and fever were the most frequent clinical manifestations, 33% of patients presented gastrointestinal symptoms. In literature gastrointestinal manifestations are irregular and nowadays it is described in 20% of the cases (6, 14, 17). Similar to other studies, the frequent coinfection was with Mycoplasma pneumoniae and in less proportion with other respiratory viruses (15, 18, 19). In relation to that, it was described that viral respiratory infections could predispose the development of over-aggregated bacterial infections like infections with M. pneumoniae due to the alteration of respiratory mucociliary clearance and the immune system response (20). Nevertheless, M. pneumoniae infection could also precede viral infection (21).

Comparable to our findings, it has been reported in other studies that the most frequent comorbidities were neurological with 11% (65/614) and hemato-oncological 8% (45/614) (6). Nevertheless, another study found the most frequent comorbidities asthma (12.3%), followed by anemia of sickle cells (7%) (20). Many of these comorbidities are related to congenital pathologies unlike adults. Regarding the severity of the disease, in this study 68.2% of PICU patients presented a comorbidity which is a high percentage which relates with other studies that report more than 80% of ICU patients hospitalized with comorbidities (4).

An increase of CPR and Dimer-D was fundamentally observed in this study. In accordance to findings in literature, a variety of laboratory alterations have been described in COVID-19, especially in inflammatory serum markers (6, 15, 23, 24) a study found high levels of CPR, procalcitonin, interleukin-6, ferritin and Dimer-D in relation to severity (23). Our findings could have been explained by the elevated proportion of patients with mild symptoms, moderate and because only initial tests were included in the analysis.

In relation to MIS-C cases, 6 patients with MIS-C diagnostic were found and 2 required PICU admission similar to what was described in another Peruvian report (⅛) (24). Meanwhile, in another study MIS-C cases represented 5 time more probabilities of PICU admission, however, no MIS-C patient died (6). In both studies all patients had a favorable evolution. In comparison, in a MIS-C systematic review in children and adolescents, it was described that MIS-C seems to be a condition of major severity with 38% (531/783) of cases requiring PICU admission (25).

Antibiotics prescription was high despite few microbiological findings. In the literature, a meta-analysis described in both adults and children that 71.3% had antibiotic prescription even though bacterial coinfection prevalence and COVID-19 was low and secondary bacterial coinfection in patients was 14% (26). These results suggest that antibiotics should be prescribed under a clear clinical suspicion of bacterial infection in patients with COVID-19.

In this study, hydroxychloroquine was less used and immunoglobulin use was reserved fundamentally for patients with MIS-C criteria. Moreover, in a systematic review 7.8% of children received hydroxychloroquine, 4.1% corticosteroids and 3.1% immunoglobulin (15). Regarding hydroxychloroquine, the limited scientific evidence reported initially can explain the findings in comparison to intravenous immunoglobulin which is accepted as immunomodulatory therapy in the management of Kawasaki and MIS-C (6, 24, 27, 28).

This study has some limitations, observational retrospective information was presented. A limitation was the absence of complete baseline tests proposed for COVID-19, partly explained by the diversity of clinical criteria for the request. Finally, for the diagnosis of MIS-C, not all patients underwent a serological test for SARS-CoV 2, and in the present study, only those who had a positive confirmatory test were included.

As a conclusion, SARS CoV-2 infection in children presents mild and moderate clinical manifestations. A great proportion of patients presented comorbidities, being higher in PICU admitted and deceased patients.

## Data Availability

The data that support the findings of this study are available from the corresponding author upon reasonable request.

## Acknowledgment

We would like to thank all services and offices of the INSN-SB for the support and facilities provided during the development of this study.

