## Supplemental File1 for "Clinical characteristics of children with COVID-19 admitted in a tertiary referral center in Perú"

**COVID-19 Working group**


- Sandra Shiu Sanabria
  https://orcid.org/0000-0002-1223-1960
- Yuliana del Pilar Bernal Fung
  https://orcid.org/0000-0003-4870-7315
- Jorge Ernesto Yoshihiro Nako Fuentes
  https://orcid.org/0000-0002-0277-972X
- Maria Elizabeth Llanos Cruz
  https://orcid.org/0000-0002-5554-0667
- Fernando Abraham Ledesma Cangahuala
  https://orcid.org/0000-0002-0979-9491
- Teresa Alegre Tuesta
  https://orcid.org/0000-0003-1745-4570
- Luz Katherine Junes Lopez
  https://orcid.org/0000-0001-9964-9763
- Elizabeth Karon Saavedra Rodriguez
  https://orcid.org/0000-0003-0224-6832
- Aldo Marcos Roberto Paz Marchena
  https://orcid.org/0000-0002-1769-1149
- Ronald Abel Pérez Apaza
